# Immunogenicity and reactogenicity of the adjuvanted respiratory syncytial virus vaccine in patients with chronic kidney disease

**DOI:** 10.64898/2025.12.02.25341408

**Authors:** Richard Radun, Saskia Bronder, Amina Abu-Omar, Danilo Fliser, Martina Sester, David Schmit

**Affiliations:** Department of Internal Medicine IV, Saarland University Medical Center, Campus Homburg, Germany; Department of Transplant and Infection Immunology, PharmaScienceHub (PSH), Saarland University, Homburg, Germany; Center for Gender-specific Biology and Medicine (CGBM), Saarland University, Homburg, Germany

**Keywords:** Respiratory syncytial virus, RSVpreF3-vaccination, chronic kidney disease, T-cells, immunoglobulins

## Abstract

**Background:** Chronic kidney disease (CKD) contributes to global mortality and morbidity, also due to infectious complications resulting from immune system dysregulation. Recently, respiratory syncytial virus (RSV) vaccines based on the prefusion F (preF) glycoprotein have been licensed for prevention of severe disease in elderly and in patients with comorbidities, but data on immunogenicity in patients with CKD is scarce.

**Methods:** We characterized humoral and cellular immunogenicity of 75 patients with CKD stages G3a to G5d both before and 14 (IQR 2) days after vaccination with an adjuvanted protein-based RSVpreF3-vaccine using ELISA and flow cytometry. Data on adverse events were collected through a self-reported questionnaire.

**Results:** Vaccination led to a significant induction of RSV-specific CD4 T-cells (p<0.0001) and the increase did not differ between the CKD-stages. CD8 T-cells were not specifically induced. Despite high seroprevalence prior to vaccination, quantitative levels of RSV-specific immunoglobulins IgG and F protein-specific IgG were significantly induced upon vaccination (both p<0.0001), with a less pronounced increase in patients with advanced CKD. Urinary albumin-creatinine-ratio (UACR) was shown to be predictive of vaccine response in a multivariate regression model using age, serum creatinine and urea as covariates (p=0.035) The vaccine was well tolerated with mostly transient adverse events at the injection site.

**Conclusions:** RSV-vaccination led to a robust CD4 T-cell and humoral response in patients with CKD with less pronounced effects in those with high-grade proteinuria. Long-term data on immunogenicity and correlation with clinical outcomes are warranted to define optimal vaccination strategies.

**Key learning points:**

- **What was known:** There is limited data on RSV-vaccine immunogenicity in patients with CKD despite them being a high-risk group for developing severe RSV-associated lower respiratory tract infection
- **This study adds:** Administration of the AS01_E_-adjuvanted RSVpreF3-vaccine significantly induces RSV-specific IgG and CD 4 T-cells in patients with CKD, including those on hemodialysis and under medical immunosuppression. Humoral vaccine response is reduced in advanced CKD and inversely correlates with UACR
- **Potential impact:** RSV-vaccination is safe and immunogenic in patients with CKD, supporting the integration of RSV-vaccination into routine preventive care for the CKD-population

## Introduction

Respiratory syncytial virus (RSV) infection poses a major risk for developing virus-associated lower respiratory tract infections. In particular, high-risk groups including elderly and chronically ill individuals such as patients with chronic kidney disease (CKD) show high rates of RSV-associated complications with a global annual estimate of over 5 million hospitalizations and 100’000 deaths worldwide (1). Recently, RSV-vaccination strategies using protein-based or mRNA-based vaccine platforms have been licensed for elderly and individuals with comorbidities including patients with CKD (2). Two international, randomized, placebo-controlled phase III trials using an AS01_E_-adjuvanted RSVpreF3-vaccine and the bivalent non-adjuvanted RSVpreF-vaccine have shown high vaccine efficacy of 94.1% and 85.7%, respectively, in preventing severe RSV-associated lower respiratory tract infection over the course of one RSV season in patients above 60 years of age without relevant safety concerns (3, 4).

CKD affects approximately 10% of the global population (5) and entails various facets of immunodeficiency. Innate and adaptive immunity are known to be impaired in uremia, and hemodialysis treatment as well as medical maintenance immunosuppression may further alter immune cell phenotype, antigen presentation and cytokine response. Reduced vaccine responsiveness in patients with CKD has been demonstrated for influenza, hepatitis B, and pneumococcal vaccines (6-11), where patients with advanced CKD and those receiving dialysis mount weaker and shorter-lived immune responses compared to the general population (12). Despite the recent surge in RSV-vaccine research, there is limited data on immunogenicity and reactogenicity of RSV-vaccination in patients with CKD.

We therefore explored the cellular and humoral immunogenicity of RSV-vaccination in patients with CKD KDIGO-stages G3-G5. A subset of patients received maintenance immunosuppression or intermittent hemodialysis treatment. To this end, we characterized RSV-specific T-cells and immunoglobulins in 75 patients with various CKD-etiologies receiving the AS01_E_-adjuvanted RSVpreF3-vaccine and assessed reactogenicity using a self-reported standardized questionnaire in this prospective cohort study.

## Material and Methods

### Study design and subjects

From January 2025 to May 2025, patients with CKD (KDIGO-stages G3a to G5d) were prospectively enrolled at the Saarland University Medical Center in Homburg, Germany. All patients received a single dose of the protein-based AS01_E_-adjuvanted vaccine RSVPreF3 (“Arexvy”, GSK) intramuscularly according to national vaccination recommendations. Further details on collected demographic and clinical data and routine laboratory parameters are given in the supplementary methods. The study was performed in adherence with the declaration of Helsinki and approved by the ethics committee of the Saarland Medical Association (reference 99/24), and written informed consent was obtained from all individuals.

### Quantification and characterization of RSV-specific CD4 and CD8 T-cells

RSV-specific CD4 and CD8 T-cells were quantified and characterized using flow cytometry after a 6-hour stimulation of whole blood samples as previously described (13, 14). A detailed description of the procedure is found in the supplementary methods including antibodies used for staining (**table S1**), and a gating strategy for flow-cytometric analyses (**figure S1)**.

### Determination of RSV-specific antibodies

Specific IgG and IgA antibodies towards pan-RSV and RSV-F (fusion protein) specific IgG were measured as described in the supplementary methods.

### Statistical analysis

All statistical analyses were performed using GraphPad Prism 10.6 software (GraphPad, San Diego. CA, USA). Statistical comparisons were conducted using non-parametric tests in case of non-normality of data. The Mann–Whitney U-test was applied for comparisons between two independent groups, while comparisons of more than two unpaired groups were performed using the Kruskal–Wallis H test followed by Dunn’s post hoc test in case of reaching significance. For paired non-parametric data, the Wilcoxon signed-rank test was employed. Linear correlations between immunological and clinical data were assessed using Spearman’s rank correlation coefficient, and multivariate logistic regression analysis was performed using R 4.5.1 software to determine predictors of immunogenicity. Parametric data were analyzed with an unpaired t-test, whereas nominal and dichotomous variables were compared using the chi-squared test. A two-sided significance level of p<0.05 was considered statistically significant.

## Results

### Study population

In this observational study, 75 patients with CKD were recruited with 17 to 21 patients in each CKD-stage from G3a to G5d. Fifteen out of 19 patients within the CKD-G5 cohort were on intermittent maintenance hemodialysis. Relevant demographic and clinical features including kidney disease, comorbidities, immunosuppression, laboratory parameters and differential blood counts are shown in Table 1. The mean age of all patients was 74.7 years with no significant differences between groups and sex distribution was comparable across all groups. The median interval between vaccination and follow-up visit was 14 (IQR 2) days and did not differ between groups. There were no significant differences in the automated differential blood counts with regards to total leukocyte number among the patients except for a non-significant trend towards a higher number of monocytes in patients with CKD-G5/G5d. Serum levels of creatinine, urea and intact parathyroid hormone and UACR increased progressively with more advanced CKD. Conversely, hemoglobin levels and estimated glomerular filtration rate (eGFR) decreased. Eleven patients received B-cell or plasma cell-depleting therapy within the past 6 months (e.g. rituximab), ongoing treatment with calcineurin inhibitors, antimetabolites, glucocorticoids or complement inhibitors. The prevalence of diabetes mellitus was similar in all groups whereas an autoimmune disease affecting the kidneys was more frequent in the group with advanced CKD. These included vasculitis, IgA nephropathy, membranous or immune complex glomerulonephritis as well as renal amyloidosis.

**Table 1.**
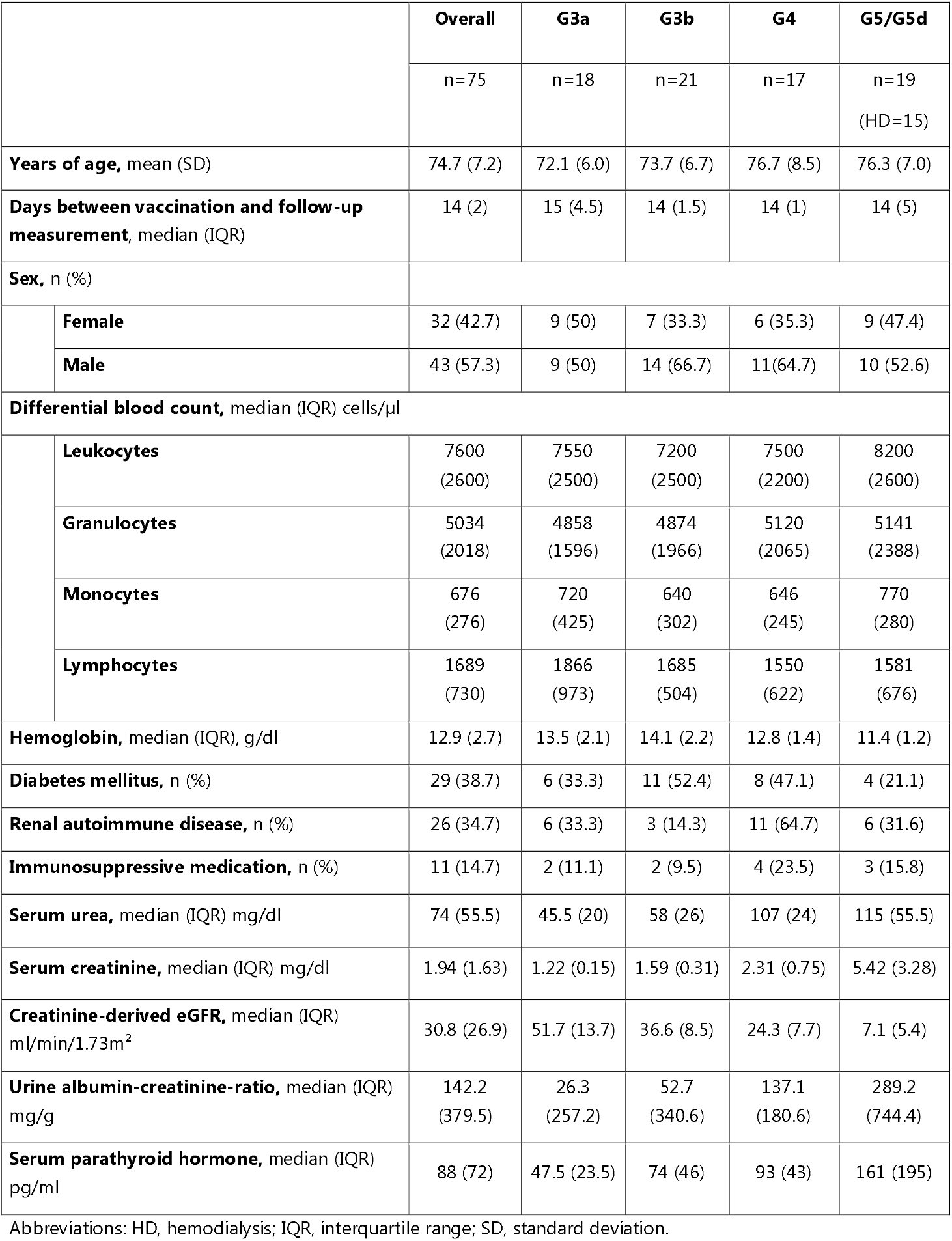
Clinical and demographic data on CKD-study patients.

|  |  | **Overall** | **G3a** | **G3b** | **G4** | **G5/G5d** |
| --- | --- | --- | --- | --- | --- | --- |
|  |  | n=75 | n=18 | n=21 | n=17 | n=19<br>(HD=15) |
| **Years of age**, mean (SD) |  | 74.7 (7.2) | 72.1 (6.0) | 73.7 (6.7) | 76.7 (8.5) | 76.3 (7.0) |
| **Days between vaccination and follow-up measurement**, median (IQR) |  | 14 (2) | 15 (4.5) | 14 (1.5) | 14 (1) | 14 (5) |
| **Sex**, n (%) |  |  |  |  |  |  |
|  | **Female** | 32 (42.7) | 9 (50) | 7 (33.3) | 6 (35.3) | 9 (47.4) |
|  | **Male** | 43 (57.3) | 9 (50) | 14 (66.7) | 11(64.7) | 10 (52.6) |
| **Differential blood count**, median (IQR) cells/μl | | | | | | |
|  | **Leukocytes** | 7600<br>(2600) | 7550<br>(2500) | 7200<br>(2500) | 7500<br>(2200) | 8200<br>(2600) |
|  | **Granulocytes** | 5034<br>(2018) | 4858<br>(1596) | 4874<br>(1966) | 5120<br>(2065) | 5141<br>(2388) |
|  | **Monocytes** | 676<br>(276) | 720<br>(425) | 640<br>(302) | 646<br>(245) | 770<br>(280) |
|  | **Lymphocytes** | 1689<br>(730) | 1866<br>(973) | 1685<br>(504) | 1550<br>(622) | 1581<br>(676) |
| **Hemoglobin**, median (IQR), g/dl |  | 12.9 (2.7) | 13.5 (2.1) | 14.1 (2.2) | 12.8 (1.4) | 11.4 (1.2) |
| **Diabetes mellitus**, n (%) |  | 29 (38.7) | 6 (33.3) | 11 (52.4) | 8 (47.1) | 4 (21.1) |
| **Renal autoimmune disease**, n (%) |  | 26 (34.7) | 6 (33.3) | 3 (14.3) | 11 (64.7) | 6 (31.6) |
| **Immunosuppressive medication**, n (%) |  | 11 (14.7) | 2 (11.1) | 2 (9.5) | 4 (23.5) | 3 (15.8) |
| **Serum urea**, median (IQR) mg/dl |  | 74 (55.5) | 45.5 (20) | 58 (26) | 107 (24) | 115 (55.5) |
| **Serum creatinine**, median (IQR) mg/dl |  | 1.94 (1.63) | 1.22 (0.15) | 1.59 (0.31) | 2.31 (0.75) | 5.42 (3.28) |
| **Creatinine-derived eGFR**, median (IQR)<br>ml/min/1.73m<sup>2</sup> |  | 30.8 (26.9) | 51.7 (13.7) | 36.6 (8.5) | 24.3 (7.7) | 7.1 (5.4) |
| **Urine albumin-creatinine-ratio**, median (IQR)<br>mg/g |  | 142.2<br>(379.5) | 26.3<br>(257.2) | 52.7<br>(340.6) | 137.1<br>(180.6) | 289.2<br>(744.4) |
| **Serum parathyroid hormone**, median (IQR)<br>pg/ml |  | 88 (72) | 47.5 (23.5) | 74 (46) | 93 (43) | 161 (195) |

Abbreviations: HD, hemodialysis; IQR, interquartile range; SD, standard deviation.

### Vaccine-induced humoral and cellular immunity in patients with CKD

For characterization of humoral immunogenicity of the RSVPreF3-AS01_E_-vaccine, RSV-specific IgG, IgA and F-IgG antibodies were measured. After a median follow-up of 14 days post vaccination, median pan-RSV-specific IgG levels showed a significant increase from 110 (IQR 41) RU/ml to 128 (IQR 32) RU/ml (p<0.0001, geometric mean 1.21-fold Figure 1A). Seropositivity defined as the percentage of individuals with an IgG level above 18 RU/ml was high with 100% before and after vaccination (Figure 1A). Likewise, vaccine RSV-F protein-specific IgG antibodies were detectable already prior to vaccination, and showed a significant increase from 152 (IQR 198) U/ml to 1305 (IQR 2669) U/ml (5.99-fold, p<0.0001). Pan-RSV-specific IgA levels were measured in a subgroup of patients. Despite low overall levels IgA levels showed a significant induction (1.51-fold, p=0.030, Figure 1A).

**Figure 1:**
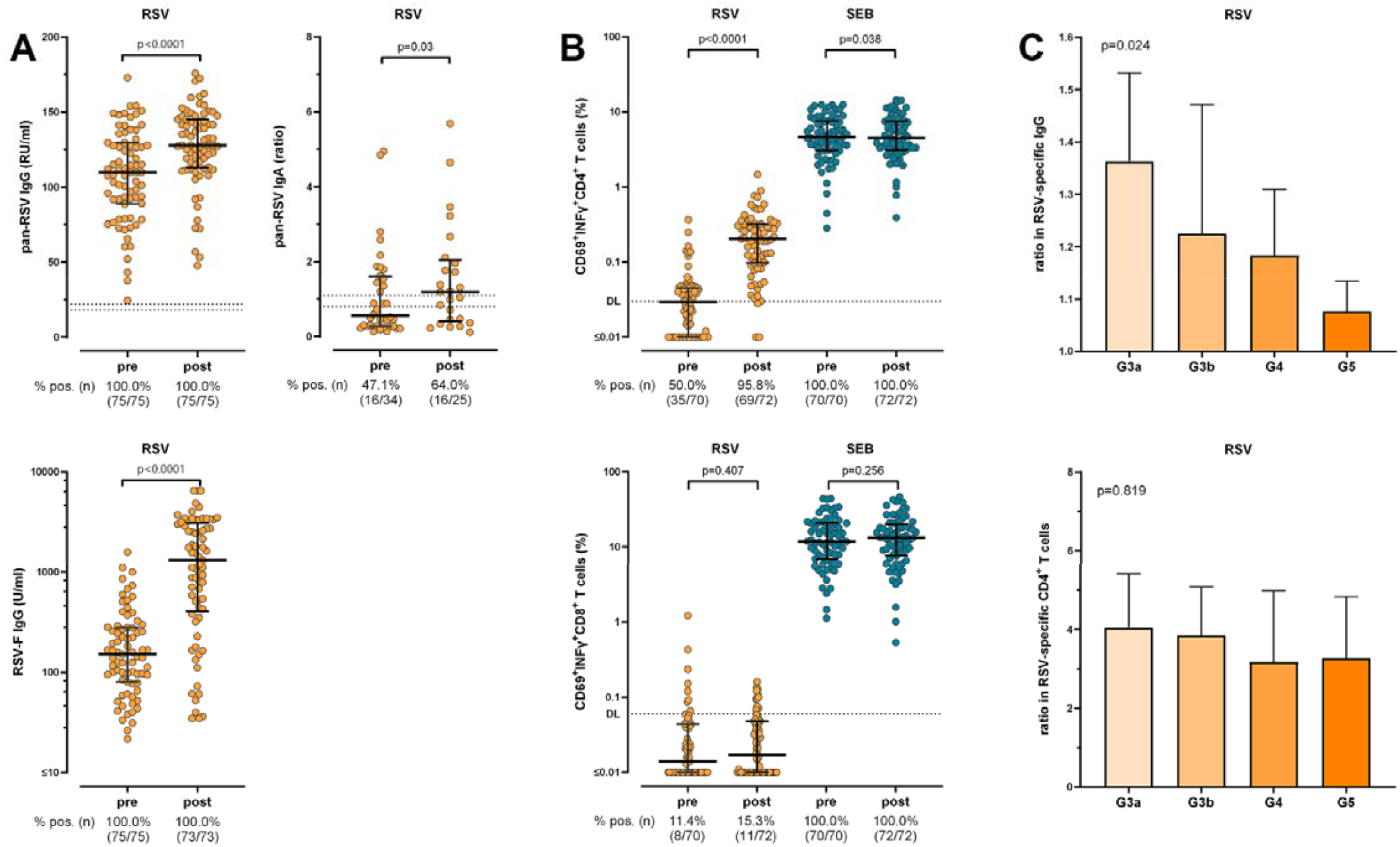
RSV-specific humoral and cellular vaccine response in patients with CKD. **(A)** Relative concentrations of RSV-specific IgG, IgA and prefusion F-specific IgG before (pre) and 14 days after (post) vaccination with the adjuvanted RSVpreF3-vaccine (Arexvy), including corresponding percentages of seropositivity. The dashed line indicates the manufacturer’s cut-off for seropositivity (if available). Dots represent individual patients; lines represent medians and interquartile ranges. **(B)** Percentages of RSV-specific CD69^+^, IFNγ^+^ CD4 and CD8 T-cells before (pre) and 14 days after (post) vaccination, including polyclonal stimulation with SEB and corresponding percentages of individuals above detection limits (DL, indicated by the dashed lines). Dots represent individual patients; lines represent medians and interquartile ranges. **(C)** Relative increase in cellular and humoral vaccine response for different CKD-stages. Columns represent geometric means with 95% confidence intervals. P-values were calculated from paired values using the Wilcoxon matched pairs test, (c) p-values were calculated using the Kruskal-Wallis test. CD, cluster of differentiation; IFN, interferon; Ig, immunoglobulin; RSV, respiratory syncytial virus; SEB, *Staphylococcus aureus* enterotoxin B.

RSV-specific CD4 and CD8 T-cells were quantified based on induction of IFNγ after stimulation with RSV-derived peptides. Upon vaccination, a significant increase in the percentage of RSV-specific activated CD4 T-cells was observed (p<0.0001, Figure 1B). Before vaccination, 50% of patients showed RSV-specific CD4 T-cells above the detection limit with an increase to 96% after vaccination (Figure 1B). CD8 T-cells showed higher variability at baseline and were not significantly induced upon vaccination (Figure 1B). A pronounced CD4 and CD8 T-cell response was detected after polyclonal stimulation with SEB, which was largely unaffected by RSV-vaccination.

Differentiation of groups according to their clinical CKD-stages revealed significant differences with highest geometric mean increases in pan-RSV-specific IgG titers in the G3a patient group with a 1.36-fold induction. In stages G3b, G4, and G5 the increases were 1.23-fold, 1.18-fold, and 1.08-fold, respectively (p=0.024). Similarly, there was a numerical decline in RSV-specific CD4 T-cells with higher CKD-stages without reaching statistical significance (p=0.819, Figure 1C).

### Functional and phenotypical characterization of RSV-specific T-cells

RSV-specific CD4 and CD8 T-cells were further characterized for their capacity to produce the cytokines TNF and IL-2. As with T-cells producing IFNγ, the percentage of CD69^+^ CD4 T-cells producing TNF and IL-2 also showed a significant induction after vaccination, whereas respective CD8 T-cell populations were not induced (figure S2A). Among vaccine induced RSV-specific CD4 T-cells, the majority of cells (54.9%) were polyfunctional with the capability to produce all three cytokines simultaneously, while 31.5% were dual-cytokine positive, and 13.6% of cells produced one cytokine only (figure S2B). This pattern was distinct from that of polyclonally stimulated T-cells, where 19.5% of cells were triple positive and similar fractions produced two (43.5%) or one cytokine (37.0%). As a sign for recent antigen encounter, numerical expansion of RSV-specific CD4 T-cells after vaccination was associated with a significant upregulation of CTLA-4 as compared to pre-vaccination levels (p<0.0001). CTLA-4 expression on CD4 T-cells after polyclonal stimulation was low and was unaffected by the vaccine (p=0.304, figure S2C).

### Immunogenicity in CKD-subgroups

To further assess RSV-vaccine immunogenicity among CKD-subgroups, 15 patients on intermittent hemodialysis were compared with non-dialysis patients (Figure 2A). Baseline levels of pan-RSV-specific IgG were higher in dialysis patients (p=0.007). Levels increased significantly in both groups, but to a lesser extent in patients on hemodialysis than in non-dialysis patients (p<0.018). In contrast, baseline levels of RSV-F-specific IgG were lower in patients on dialysis, and were robustly induced in both groups without significant differences in patients with and without dialysis (p=0.469). Patients on dialysis had a higher proportion of RSV-specific CD4 T-cells above detection limit at baseline (p=0.034), which increased to a similar extent as in non-dialysis patients p=0.330, Figure 2A).

**Figure 2:**
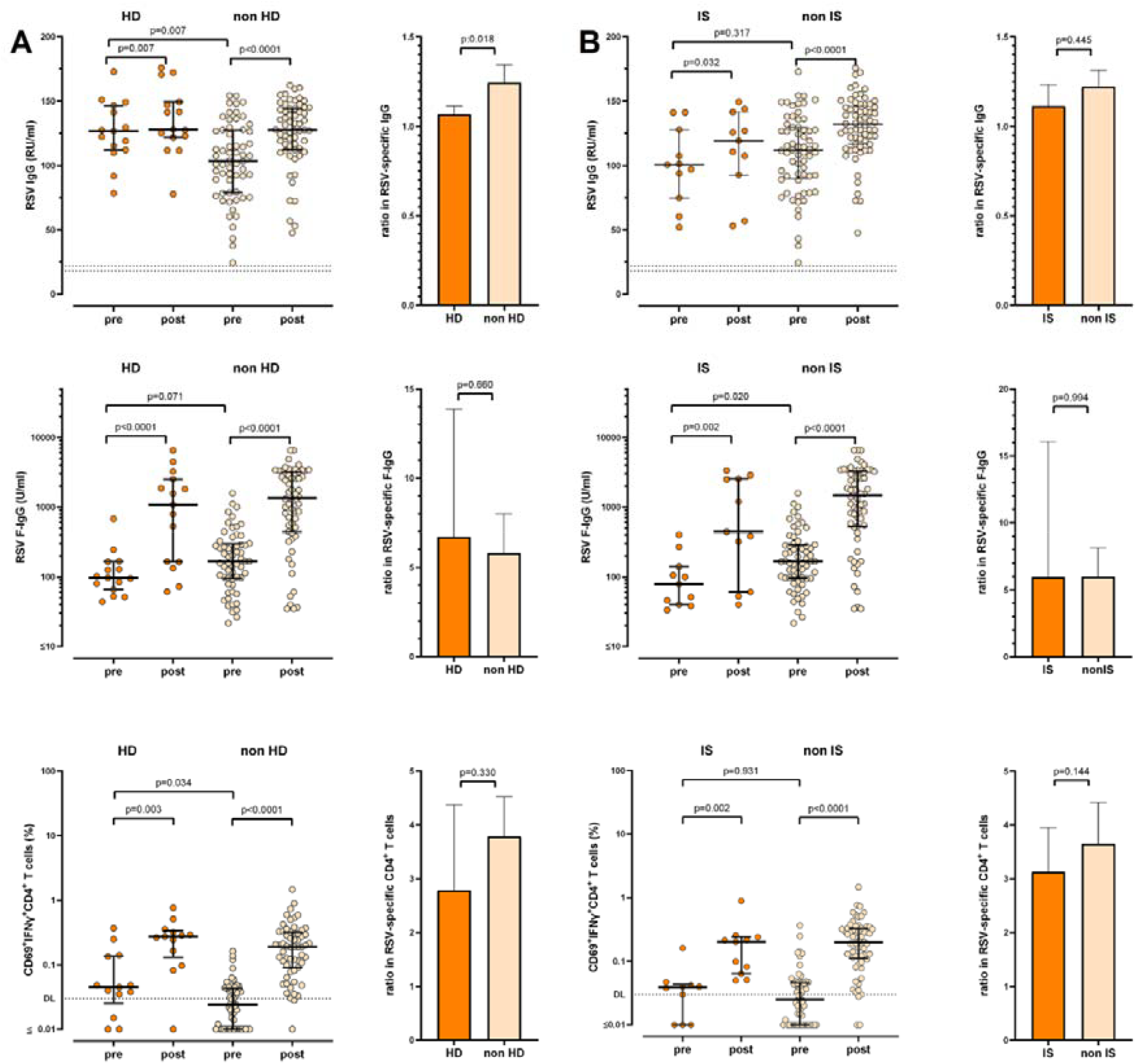
Differential cellular and humoral effects of RSV-vaccination in hemodialysis patients and immunosuppressed patients. Levels of pan-RSV-specific IgG and RSV-F-IgG, as well as percentages of RSV-specific CD4 T-cells before (pre) and 14 days after (post) vaccination in **(A)** patients on intermittent hemodialysis (HD, n=15) versus non-dialysis patients (non HD, n=60), and in **(B)** patients under medical maintenance immunosuppression (IS, n=11) versus patients without medical immunosuppression (non IS, n=64) including comparison of the fold increase upon vaccination. Dots represent individual patients; lines represent medians and interquartile ranges; columns represent geometric means with 95% confidence intervals; p-values were calculated from paired values using the Wilcoxon matched pairs test, p-values for fold-increases were calculated using the Mann-Whitney test. CD, cluster of differentiation; IFN, interferon; Ig, immunoglobulin; RSV, respiratory syncytial virus; SEB, *Staphylococcus aureus* enterotoxin B.

When comparing a subset of 11 patients with CKD receiving maintenance immunosuppressive therapy with patients without immunosuppression, both subgroups showed a significant increase in IgG antibodies (pan-RSV, F-specific) and RSV-specific CD4 T-cells. However, there was no difference in the extent of increase between the groups (Figure 2B). Finally, patients with CKD were stratified into an advanced disease cohort (eGFR <30ml/min/1,73m^2^, >G4) and less advanced cohort (G3a/b). In both groups, RSV-specific IgG were robustly induced upon vaccination with a less pronounced increase in advanced CKD (p=0.030, Figure S3). Similarly, a significant induction of RSV-F-specific IgG and RSV-specific CD4 T-cells was observed in both groups, but the extent of increase did not differ between the groups (Figure S3).

### Clinical correlations and predictors of immune response

Standardized clinical parameters associated with CKD and its progression were collected and analyzed in terms of their correlation with cellular and humoral immunogenicity. A moderate but significant inverse linear correlation was found between the relative increase in RSV-specific IgG levels and the UACR at the time of vaccination (Spearman r=-0.261, p=0.030, Figure 3A). A similar inverse relation was found between the increase in RSV-specific CD4 T-cells and UACR or CRP as marker of systemic inflammation, but this did not reach statistical significance (UACR: Spearman r=-0.176, p=0.614, Figure 3A; CRP: r=-0.091, p=0.441, Figure S4A). eGFR correlated positively with RSV-IgG vaccine responses (Spearman r=0.301, p=0.03, Figure 3B), in line with an inverse correlation of serum urea with RSV-IgG levels (Spearman r=-0.265, p=0.024, Figure S4A), respectively. In a multivariate logistic regression model a significant relationship between UACR and predicted vaccine responder probability defined as having an increase in both RSV-specific IgG and CD4 T-cells was observed, showing significant decline of responder probability with high UACR levels (p=0.035, Figure 3C). In contrast, when CD4 T-cell or IgG responses were analyzed individually, no significant association with log-transformed UACR were observed despite a non-significant trend for humoral vaccine response (p=0.412 for CD4 responses and p=0.077 for IgG responses, Figure S4B).

**Figure 3:**
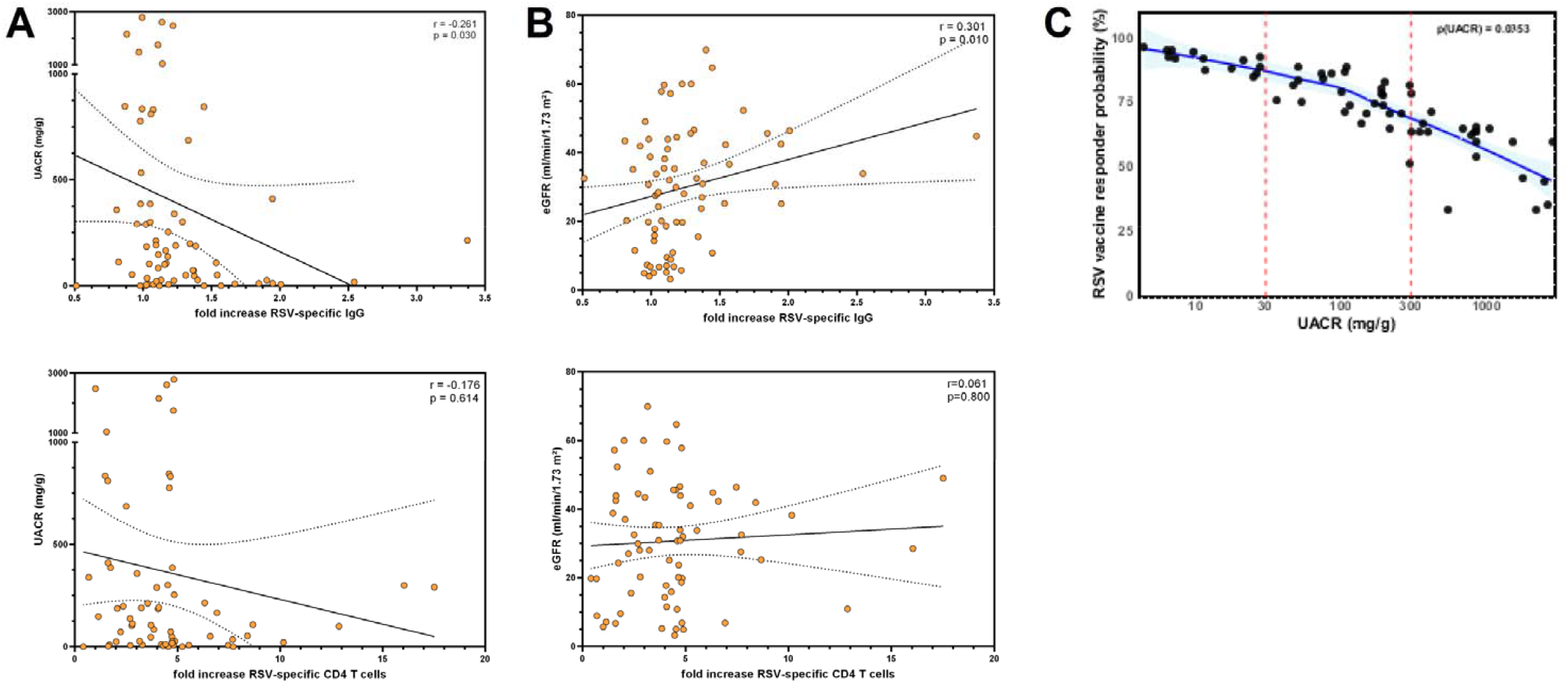
Correlations of humoral and cellular vaccine response with clinical CKD-parameters pre vaccination. **(A)** Linear inverse correlations between the fold increase in RSV-specific IgG or RSV-specific CD4 T-cells and the urine albumin-to-creatinine ratio (UACR). Dots represent individual patients, orange-colored dots denote patients with immunosuppression. Lines correspond to the linear regression line with overlaid confidence intervals. **(B)** Linear correlation between fold increase in RSV-specific IgG and creatinine-derived eGFR. Dots represent individual patients. Lines correspond to the linear regression line with overlaid confidence intervals. **(C)** Predicted probability of being a vaccine responder defined as having an increase in both RSV-specific IgG and CD4 T-cells, based on a multivariate logistic regression model including log-transformed UACR as predictor and eGFR, serum urea and age as covariates. The blue line represents a locally weighted scatterplot smoothing (LOESS) curve with other covariates set at the median, the light blue area displays the 95% confidence interval. Dots represent individual patients. CD, cluster of differentiation; CRP, C-reactive protein; eGFR, estimated glomerular filtration rate; Ig, immunoglobulin; RSV, respiratory syncytial virus; UACR, urine albumin-to-creatinine ratio.

### Reactogenicity of RSV-vaccination in patients with CKD

Reactogenicity was assessed in all patients using a questionnaire on self-reported adverse events within the first 7 days. No adverse events were reported by 33% of patients, while 61% reported transient local reactions such as pain or swelling at the injection site. Twenty-three percent of patients stated systemic symptoms, most commonly arthralgia, myalgia, fatigue or elevated body temperature (Figures 4A and 4B). None of the symptoms persisted beyond two weeks or led to hospitalization. To evaluate a potential association between reactogenicity and cellular immune response, the relative increase in RSV-specific CD4 T-cells was compared between patients with and without adverse events. However, no significant differences were observed (Figure 4C).

**Figure 4:**
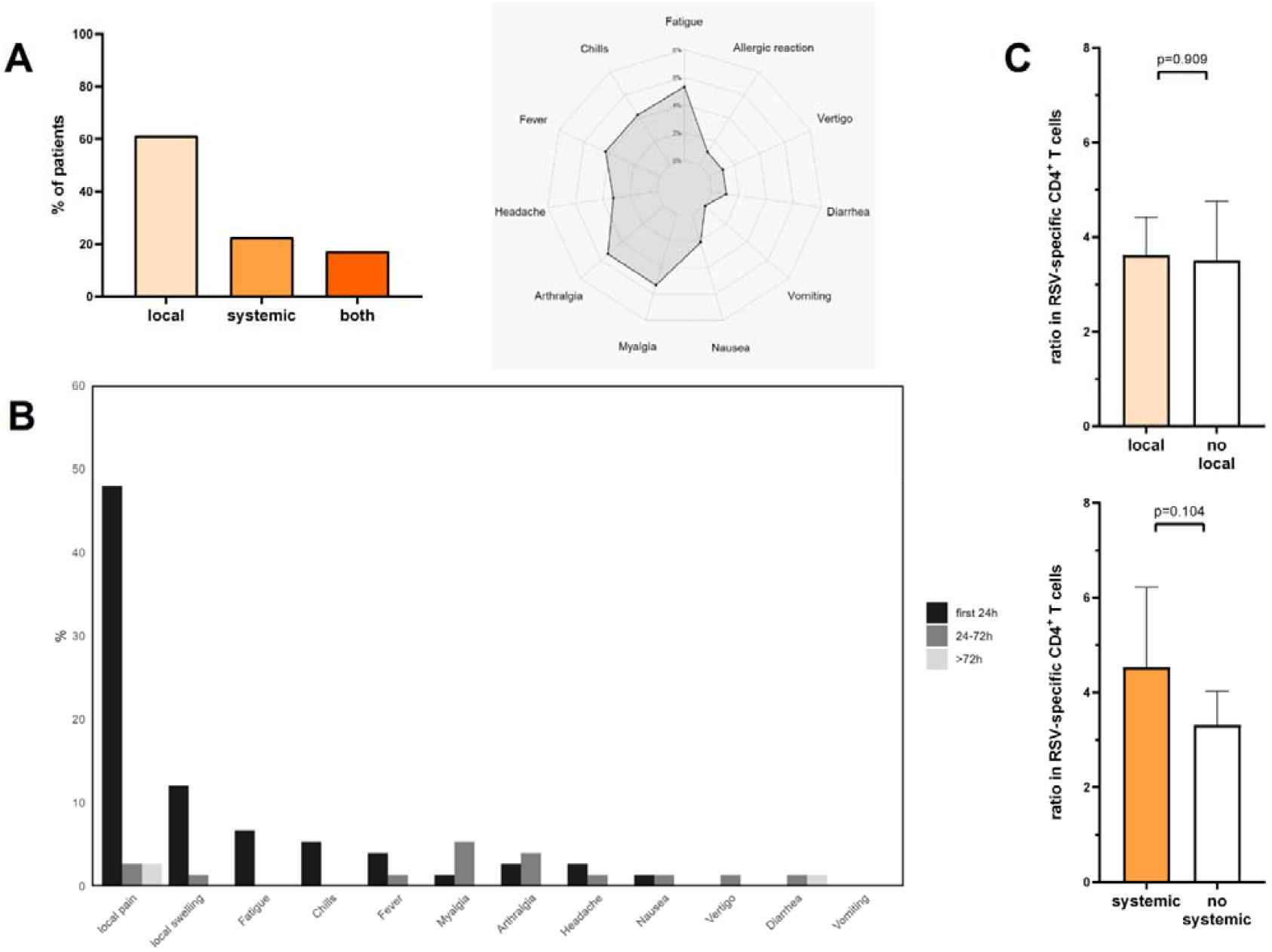
Reactogenicity of RSV-vaccination in patients with CKD. **(A)** Distribution of self-reported adverse events at different time points after vaccination. Bars represent percentage of patients. Radar plot illustrating systemic adverse events. Line represents percentage of patients with self-reported events. **(B)** Temporal distribution of self-reported adverse events within the first week after vaccination. Groups are stratified as first occurrence of symptoms within 24 hours, between 24 and 72 hours or between 72 hours and one week after vaccination. Bars represent percentage of patients. **(C)** Fold increase in induction of RSV-specific CD4 T-cells for patients with or without local or systemic reactions. Columns represent geometric means with 95% confidence intervals. Columns represent geometric means with 95% confidence intervals; p-values were calculated using the Mann-Whitney test. CD, cluster of differentiation; RSV, respiratory syncytial virus;

## Discussion

Due to a lack of specific antiviral therapies and risk for severe disease course in high-risk groups, RSV-vaccination poses a readily available preventive strategy for reducing RSV-associated morbidity and mortality. However, data on immunogenicity and reactogenicity of RSV-vaccination in patients with CKD is still scarce (2). In line with global endemicity of RSV, we found a high rate of baseline seropositivity across all CKD-stages. Upon vaccination with a single dose of the adjuvanted RSVpreF3-vaccine, a significant increase in pan-RSV IgG and IgA, RSV-F-specific IgG as well as RSV-specific CD4 T-cells was observed. There was no induction of RSV-specific CD8 T-cells. The magnitude in the induction of humoral immune response was lower in patients on intermittent hemodialysis, while patients under maintenance medical immunosuppression showed similar cellular and humoral vaccine response as non-immunosuppressed individuals. An advanced CKD-stage was associated with a lower humoral immunogenicity, and higher UACR was related to a lower vaccination response in a logistic regression model. RSV-vaccination was well tolerated and the majority of patients reported only minor or transient local adverse events.

The study cohort had a mean age of 74.7 years and a median eGFR of 30 ml/min/1.73m^2^, representing a high-risk population for RSV-associated lower respiratory tract infections and comparable to other published CKD-cohorts (15). Patients were evenly distributed in terms of KDIGO-stages, age, and sex, and their lymphocyte counts did not differ. Comorbidities were comparable across the groups with median prevalence of diabetes of 38.7%, kidney autoimmune disease of 34.7% and immunosuppressive therapy of 15% (16).

In line with previous reports on RSV seroprevalence, RSV-specific immunoglobulins were detected in all patients at baseline (13, 17, 18). Upon vaccination, pan-RSV-IgG, pan-RSV-IgA and RSV-F-specific IgG significantly increased together with an increase in RSV-specific CD4 T-cells as seen in other reports on immunocompromised individuals (13, 19, 20). Similarly, recent reports found seroconversion in 61% of immunosuppressed patients (21). Unlike for RSV-specific CD4 T-cells, vaccination had no immediate effect on RSV-specific CD8 T-cells. This aligns with previous findings on immunogenicity of adjuvanted RSVpreF3-vaccine and other protein-based vaccines such as the shingles-vaccine Shingrix or the COVID-19-vaccine NVX-CoV2373(22-26). This lack of effect can be attributed to the mechanism that peptides from protein-based vaccines are primarily presented through MHC class II therefore predominantly triggering a CD4 T-cell response (23).

For patients undergoing intermittent hemodialysis, induction of humoral immune response was significantly lower compared to non-dialysis patients in line with a numerically smaller increase in RSV-specific cellular immune response. Notably, this less pronounced increase in dialysis patients may be due to higher pan-RSV-specific IgG- and CD4 T-cell levels even before vaccination, which likely reflect prior exposure to RSV and underscores their increased infection risk. Despite exclusion of respiratory infection at the time of vaccination, subclinical RSV infection may have contributed to the inability to clearly distinguish vaccine-specific effects. These findings align with previous reports of impaired vaccine responses in dialysis patients, including lower antibody responses as well as faster waning of both cellular and humoral immune responses to vaccinations (6, 27, 28). Similarly, patients under medical immunosuppression exhibited a numerically smaller increase in CD4 and RSV-specific IgG vaccine response without reaching statistical significance. This likely may result from the small sample size and heterogeneity of immunosuppressive regimens, including B-cell depleting antibodies and antimetabolites which are commonly known to impair humoral vaccine response (29, 30). For the subgroup of immunosuppressed patients receiving complement inhibitors, effects of vaccine immunogenicity are poorly characterized. Preclinical studies suggest that C3b reduces complement activation and humoral response to adenovirus vectors (31), while clinical data indicate impaired antibody neutralization and altered T-cell memory development in viral infections such as influenza (32, 33). RSV-vaccination immunogenicity has previously been studied in kidney and lung transplant recipients demonstrating strong cellular and humoral vaccine induction (13, 19, 20). Future studies should address the stability of the vaccine immune response and effectiveness towards protection from RSV infection in both patients with CKD and solid organ transplant recipients.

Heterogeneity of cellular and humoral immune response prompted the question of defining predictive parameters of vaccine response. Interestingly, the increase in RSV-specific IgG but to a lesser extent of CD4 T-cells showed an inverse correlation with UACR. Furthermore, there was a significant negative inverse correlation between RSV-specific IgG with serum urea levels as well as a significant positive correlation between RSV-specific IgG and eGFR in line with impaired immune function in uremia. In a multivariate logistic regression model including UACR, eGFR, serum urea and age as covariates, the vaccine response probability defined as an increase of both RSV-specific IgG and CD4 T-cells was in part predicted by pre-vaccination UACR. This effect was mostly attributable to the humoral vaccine response. There is limited data on vaccine immunogenicity in relation to proteinuria. In a small Japanese cohort, patients with nephrotic syndrome under medical immunosuppression mounted lower spike-specific antibody responses to SARS-CoV-2 vaccination (34). In a small pediatric cohort with idiopathic nephrotic syndrome, reduced antibody titers against hepatitis B and tetanus were observed despite normal antigen-specific B-cell counts (35). A possible explanation for diminished immune response in highly proteinuric patients might be urinary loss of immunoglobulins or potentially even loss of vaccine antigen or adjuvants due to impaired filtration barrier. However, given intramuscular application and lymphatic transport to draining lymph nodes, systemic losses are likely of limited magnitude.

Vaccination using the adjuvanted RSVpreF3-vaccine was well tolerated with mostly transient and primarily local adverse events. Only a minority of patients reported systemic adverse events such as arthralgia or myalgia and no symptoms persisted beyond 7 days. This is in line with previous reports (3), including transplant recipients (13, 20). Despite some associations between cellular immune responses and systemic adverse events found for other vaccines (36), no significant differences between patients with and without adverse events were found in terms of RSV-specific cellular immune response.

Study limitations are the single-center design without randomization, placebo control or blinding. The size of individual subgroups was relatively small with heterogeneity in comorbidities and medical immunosuppression, limiting the generalizability of our findings. Furthermore, no data on neutralizing activity or durability of RSV-specific immunoglobulins were collected. As we did not systematically test for RSV infection, concurrent respiratory infection may have been present in some patients and contributed to interindividual variability of baseline- and/or vaccine-induced immunity. Finally, findings from this study are based on in vitro stimulation assays and do not directly infer clinical vaccine effectiveness in terms of reduced infection risk or disease severity.

## Conclusion

A single intramuscular dose of the adjuvanted RSVpreF3-vaccine led to a significant cellular and humoral vaccine response in all patients with CKD. This held true for both patients under immunosuppressive therapy and intermittent hemodialysis treatment despite in part lower magnitude of effect in these subgroups. UACR was identified as one of the covariates for predicting vaccine response probability. The vaccine was well tolerated with predominantly local and transient adverse events. Long-term stability of the immune responses and clinical data are warranted and careful consideration of reduced immunogenicity versus elevated infection risk and high infection-associated mortality remains essential to develop specific vaccination strategies in CKD-cohorts.

## Abbreviations

AS01_E_: adjuvant system 01_E_
CD: cluster of differentiation
CKD: chronic kidney disease
CRP: C-reactive protein
CTLA-4: cytotoxic T lymphocyte-associated protein 4
DMSO: dimethyl sulfoxide
eGFR: estimated glomerular filtration rate
ELISA: enzyme-linked immunosorbent assay
GSK: GlaxoSmithKline
HD: hemodialysis
IFNγ: interferon gamma
Ig: immunoglobulin
IL: interleukin
KDIGO: Kidney Disease: Improving Global Outcomes
MHC: major histocompatibility complex
mRNA: messenger ribonucleic acid
RSV: respiratory syncytial virus
RU: relative units
SARS-CoV-2: severe acute respiratory syndrome coronavirus 2
SEB: *Staphylococcus aureus* enterotoxin B
TNF: tumor necrosis factor
UACR: urine albumin-to-creatinine ratio

## Acknowledgements

The authors thank Ellen Becker, Claudia Noll, Caroline Abbosh, Candida Guckelmus and Rebecca Urschel for excellent technical assistance, and Fabio Lizzi and the team of the Saarland University Medical center for their support in enrolling participants. The authors also thank all participants to this study who contributed to the gain in knowledge from this project.

## Author Contributions

R.R., S.B., A.A-O., D.S., D.F., and M.S. designed the study and the experiments. S.B. performed flow cytometry measurements, R.R. performed F-specific ELISA tests, S.B. and A.A-O. performed RSV IgG and IgA ELISA tests. R.R., A.A-O., D.F., and D.S. contributed to patient recruitment, and clinical data acquisition. D.F., D.S. and M.S. supervised all parts of the study. R.R., S.B. and M.S. performed statistical analyses and wrote the manuscript. All authors approved the final version of the manuscript.

## Data availability statement

All figures and tables have associated raw data. The data that support the findings of this study are available from the corresponding authors upon request.

## References

1. Shi T, Denouel A, Tietjen AK, et al.; Global Disease Burden Estimates of Respiratory Syncytial Virus-Associated Acute Respiratory Infection in Older Adults in 2015: A Systematic Review and Meta-Analysis. J Infect Dis 2020; 222(Suppl 7):S577–S583.

2. Falman A, Schönfeld V, Flasche S, et al.; Beschluss und Wissenschaftliche Begründung zur Empfehlung der STIKO für eine Standardimpfung gegen Erkrankungen durch Respiratorische Synzytial-Viren (RSV) für Personen ≥ 75 Jahre sowie zur Indikationsimpfung von Personen im Alter von 60 bis 74 Jahren mit Risikofaktoren. Epid Bull 2024; 32:3–28.

3. Papi A, Ison MG, Langley JM, et al.; Respiratory Syncytial Virus Prefusion F Protein Vaccine in Older Adults. N Engl J Med 2023; 388(7):595–608.

4. Walsh EE, Perez Marc G, Zareba AM, et al.; Efficacy and Safety of a Bivalent RSV Prefusion F Vaccine in Older Adults. N Engl J Med 2023; 388(16):1465–1477.

5. Global Burden of Diseases Chronic Kidney Disease Collaboration; Global, regional, and national burden of chronic kidney disease, 1990-2017: a systematic analysis for the Global Burden of Disease Study 2017. Lancet 2020; 395(10225):709–733.

6. Sester U, Schmidt T, Kuhlmann MK, Gärtner BC, Uhlmann-Schiffler H, Sester M; Serial influenza-vaccination reveals impaired maintenance of specific T-cell memory in patients with end-stage renal failure. Vaccine 2013; 31(38):4111–20.

7. Robinson J; Efficacy of pneumococcal immunization in patients with renal disease--what is the data? Am J Nephrol 2004; 24(4):402–9.

8. Lacson E, Teng M, Ong J, Vienneau L, Ofsthun N, Lazarus JM; Antibody response to Engerix-B and Recombivax-HB hepatitis B vaccination in end-stage renal disease. Hemodial Int 2005; 9(4):367–75.

9. Beyer WE, Versluis DJ, Kramer P, Diderich PP, Weimar W, Masurel N; Trivalent influenza vaccine in patients on haemodialysis: impaired seroresponse with differences for A-H3N2 and A-H1N1 vaccine components. Vaccine 1987; 5(1):43–8.

10. Fraser GM, Ochana N, Fenyves D, et al.; Increasing serum creatinine and age reduce the response to hepatitis B vaccine in renal failure patients. J Hepatol 1994; 21(3):450–4.

11. Vandecasteele SJ, Ombelet S, Blumental S, Peetermans WE; The ABC of pneumococcal infections and vaccination in patients with chronic kidney disease. Clin Kidney J 2015; 8(3):318–24.

12. Girndt M, Sester U, Sester M, Kaul H, Kohler H; Impaired cellular immune function in patients with end-stage renal failure. Nephrol Dial Transplant 1999; 14(12):2807–10.

13. Bronder S, Abu-Omar A, Lennartz S, et al.; Cellular and humoral immunogenicity of respiratory syncytial virus vaccination in solid organ transplant recipients. Am J Transplant 2025.

14. Urschel R, Bronder S, Klemis V, et al.; SARS-CoV-2-specific cellular and humoral immunity after bivalent BA.4/5 COVID-19-vaccination in previously infected and non-infected individuals. Nat Commun 2024; 15(1):3077.

15. Liu P, Quinn RR, Lam NN, et al.; Progression and Regression of Chronic Kidney Disease by Age Among Adults in a Population-Based Cohort in Alberta, Canada. JAMA Netw Open 2021; 4(6):e2112828.

16. Fenta ET, Eshetu HB, Kebede N, et al.; Prevalence and predictors of chronic kidney disease among type 2 diabetic patients worldwide, systematic review and meta-analysis. Diabetol Metab Syndr 2023; 15(1):245.

17. Teodoro LI, Ovsyannikova IG, Grill DE, Poland GA, Kennedy RB; Seroprevalence of RSV antibodies in a contemporary (2022-2023) cohort of adults. Int J Infect Dis 2025; 158:107964.

18. Poniedzialek B, Majewska W, Kondratiuk K, et al.; Seroprevalence of RSV IgG Antibodies Across Age Groups in Poland After the COVID-19 Pandemic: Data from the 2023/2024 Epidemic Season. Vaccines (Basel) 2025; 13(7).

19. Havlin J, Skotnicova A, Dvorackova E, et al.; Respiratory syncytial virus prefusion F3 vaccine in lung transplant recipients elicits CD4+ T cell response in all vaccinees. Am J Transplant 2025; 25(7):1452–1460.

20. Hall VG, Alexander AA, Mavandadnejad F, et al.; Safety and immunogenicity of adjuvanted respiratory syncytial virus vaccine in high-risk transplant recipients: an interventional cohort study. Clin Microbiol Infect 2025.

21. Karaba AH, Hage C, Sengsouk I, et al.; Antibody Response to Respiratory Syncytial Virus Vaccination in Immunocompromised Persons. JAMA 2025; 333(5):429–432.

22. Leroux-Roels I, Davis MG, Steenackers K, et al.; Safety and Immunogenicity of a Respiratory Syncytial Virus Prefusion F (RSVPreF3) Candidate Vaccine in Older Adults: Phase 1/2 Randomized Clinical Trial. J Infect Dis 2023; 227(6):761–772.

23. Zhang Z, Mateus J, Coelho CH, et al.; Humoral and cellular immune memory to four COVID-19 vaccines. Cell 2022; 185(14):2434–2451 e17.

24. Schwarz TF, Hwang SJ, Ylisastigui P, et al.; Immunogenicity and Safety Following 1 Dose of AS01E-Adjuvanted Respiratory Syncytial Virus Prefusion F Protein Vaccine in Older Adults: A Phase 3 Trial. J Infect Dis 2024; 230(1):e102–e110.

25. Hielscher F, Schmidt T, Enders M, et al.; The inactivated herpes zoster vaccine HZ/su induces a varicella zoster virus specific cellular and humoral immune response in patients on dialysis. EBioMedicine 2024; 108:105335.

26. Hielscher F, Schmidt T, Klemis V, et al.; NVX-CoV2373-induced cellular and humoral immunity towards parental SARS-CoV-2 and VOCs compared to BNT162b2 and mRNA-1273-regimens. J Clin Virol 2022; 157:105321.

27. Espi M, Koppe L, Fouque D, Thaunat O; Chronic Kidney Disease-Associated Immune Dysfunctions: Impact of Protein-Bound Uremic Retention Solutes on Immune Cells. Toxins (Basel) 2020; 12(5).

28. Girndt M, Sester M, Sester U, Kaul H, Köhler H; Defective expression of B7-2 (CD86) on monocytes of dialysis patients correlates to the uremia-associated immune defect. Kidney Int 2001; 59(4):1382–9.

29. van der Kolk LE, Baars JW, Prins MH, van Oers MH; Rituximab treatment results in impaired secondary humoral immune responsiveness. Blood 2002; 100(6):2257–9.

30. Stumpf J, Siepmann T, Lindner T, et al.; Humoral and cellular immunity to SARS-CoV-2 vaccination in renal transplant versus dialysis patients: A prospective, multicenter observational study using mRNA-1273 or BNT162b2 mRNA vaccine. Lancet Reg Health Eur 2021:100178.

31. Mellors J, Tipton T, Longet S, Carroll M; Viral Evasion of the Complement System and Its Importance for Vaccines and Therapeutics. Front Immunol 2020; 11:1450.

32. Fernandez Gonzalez S, Jayasekera JP, Carroll MC; Complement and natural antibody are required in the long-term memory response to influenza virus. Vaccine 2008; 26 Suppl 8:I86–93.

33. Kopf M, Abel B, Gallimore A, Carroll M, Bachmann MF; Complement component C3 promotes T-cell priming and lung migration to control acute influenza virus infection. Nat Med 2002; 8(4):373–8.

34. Colucci M, Piano Mortari E, Zotta F, et al.; Evaluation of Immune and Vaccine Competence in Steroid-Sensitive Nephrotic Syndrome Pediatric Patients. Front Immunol 2021; 12:602826.

35. Kamei K, Ogura M, Sato M, et al.; Immunogenicity and safety of SARS-CoV-2 mRNA vaccine in patients with nephrotic syndrome receiving immunosuppressive agents. Pediatr Nephrol 2023; 38(4):1099–1106.

36. Schmidt T, Klemis V, Schub D, et al.; Cellular immunity predominates over humoral immunity after homologous and heterologous mRNA and vector-based COVID-19 vaccine regimens in solid organ transplant recipients. Am J Transplant 2021; 21(12):3990–4002.

