## Supplementary information for "Immunogenicity and reactogenicity of the adjuvanted respiratory syncytial virus vaccine in patients with chronic kidney disease"

### Supplementary material

This supplement contains supplementary methods as well as one supplementary **table S1** and four supplementary **figures S1-S4**.

### Supplementary methods

#### Study design, subjects, and clinical data

Demographic and clinical data including age, sex, comorbidities, underlying kidney disease and medication including immunosuppression were collected. Routine laboratory parameters, such as serum creatinine, urea, C-reactive protein (CRP), and urine albumin-to-creatinine ratio (UACR), were measured as part of standard clinical outpatient assessment. Heparinized blood samples were drawn before and at least two weeks after vaccination to characterize RSV-specific T-cell and immunoglobulin responses. Data were compared across KDIGO stages as well as after subdivision in patients with immunosuppression or patients receiving intermittent hemodialysis and their respective controls. Data on reactogenicity was gathered by self-reporting using a standardized questionnaire for local and systemic adverse events within the first seven days after vaccination.

#### Quantification and characterization of RSV-specific CD4 and CD8 T-cells

RSV-specific CD4 and CD8 T-cells were quantified and characterized using flow cytometry after a 6-hour stimulation of heparinized whole blood samples as previously described (1, 2). In brief, whole blood was stimulated in the presence of co-stimulatory antibodies against CD28 and CD49d (clone L293 and clone 9F10, 1 μg/ml each) with 2 µg/ml overlapping peptides spanning glycoprotein F0 (61%), matrix (11%) and nucleoprotein (11%) of RSV (PM-pan-RSVselect-1, jpt Berlin, Germany). In addition, stimulation with 0.64% DMSO served as a negative control, and stimulation with 2.5 μg/ml *Staphylococcus aureus* enterotoxin B (SEB; Sigma) served as positive control and as RSV-non-specific stimulus. After stimulation, cells were immunostained using anti-CD4, anti-CD8, anti-CD69, anti-IFNγ, anti-IL-2, anti-TNF, and anti-CTLA-4, and analyzed using flow cytometry (BD FACS Canto II and FACSDiva software 6.1.3., antibodies specified in **table S1**). The activation marker CD69 was used in combination with cytokines to identify RSV-specific and SEB-reactive CD4 and CD8 T-cells. Levels of CD4 and CD8 T-cells after negative control stimulation was subtracted from the RSV-specific levels and the detection limit was set at 0.03% and 0.06% of reactive CD4 and CD8 T-cells, respectively, as defined previously (1, 2). The fold change in RSV-specific CD4 T-cell immunity before and after vaccination was calculated as a ratio between post and pre-vaccination values; to avoid division by 0, the value 0.03% was added to each percentage of RSV-specific CD4 T-cells prior to division as described before (1). T-cell functionality was characterized by analyzing the co-expression of cytokines as well as the cytotoxic T-lymphocyte-associated Protein 4 (CTLA-4). The gating strategy is shown in **figure S1**.

#### Determination of RSV-specific antibodies

Specific IgG and IgA antibodies towards pan-RSV were measured using enzyme-linked immunosorbent assay kits (ELISA, anti-RSV-IgG and anti-RSV-IgA) according to the manufacturer’s instructions (Euroimmun, Lübeck, Germany). RSV-F (fusion protein) specific IgG were quantified using an ELISA according to the manufacturer’s instructions (Human Anti-RSV F protein IgG, Alpha Diagnostic International, Texas, USA). Pan-RSV-specific IgG levels (RU/ml) were categorized into negative for values <16, intermediate for values ≥16 and <22 or positive for values ≥22. The fold change between values before and after vaccination was calculated as the ratio between post- and pre-vaccination levels. IgA levels were calculated as ratios defined as the extinction of the patient sample divided by the extinction of a calibrator serum. The corresponding ratios were expressed as negative with values <0.8, intermediate with values ≥0.8 and <1.1 and positive with values ≥1.1. RSV-F-specific IgG antibodies expressed as U/ml.

#### Supplementary references

1.	Bronder S, Abu-Omar A, Lennartz S, et al.; Cellular and humoral immunogenicity of respiratory syncytial virus vaccination in solid organ transplant recipients. Am J Transplant 2025.

2.	Urschel R, Bronder S, Klemis V, et al.; SARS-CoV-2-specific cellular and humoral immunity after bivalent BA.4/5 COVID-19-vaccination in previously infected and non-infected individuals. Nat Commun 2024; 15(1):3077.

### Supplementary table

#### Supplementary table S1: Antibodies used for flow cytometric analyses

| Antigen | Conjugate | Clone | Isotype | Working concentration^#^ | Catalogue number | RRID^1^ |
| --- | --- | --- | --- | --- | --- | --- |
| CD4 | APC-H7 | SK3 | IgG κ | 1:33.3 | 641398 | AB_1645732 |
| CD8 | PerCP | SK1 | IgG1 | 1:12.5 | 345774 | AB_2868802 |
| CD152 (CTLA-4) | APC | BNI3 | IgG2a κ | 1:25 | 555855 | AB_398615 |
| CD28* |  | L293 | IgG1 κ | 1µg/ml | 348040 | AB_400367 |
| CD49d* |  | 9F10 | IgG1 κ | 1µg/ml | 555501 | AB_396068 |
| CD69 | PE-Cy7 | L78 | IgG1 κ | 1:50 | 335792 | AB_1937286 |
| IFNγ | FITC | 4S.B3 | IgG1 κ | 1:100 | 554551 | AB_395473 |
| IL-2 | PE | MQ1-17H12 | IgG2a | 1:25.5 | 559334 | AB_397231 |
| TNF | V450 | MAb11 | IgG1 κ | 1:50 | 561311 | AB_10646031 |

^1^RRID, Research Resource ID; all antibodies are mouse anti-human except anti-IL-2 which is rat anti-human; all antibodies were separately titrated in house to evaluate best concentrations for discrimination of negative and positive cell populations; panels including defined concentration of antibodies were tested together to confirm that identified antibody concentrations are also sufficient when compensations are required; ^#^The concentration of the antibodies used for antigen-specific stimulation was indicated as a concentration in a final volume of 50µl staining solution; *non-conjugated costimulatory antibodies used during antigen-specific stimulation; all antibodies from BD Biosciences, Heidelberg, Germany.

### Supplementary figures

#### Supplementary figure S1

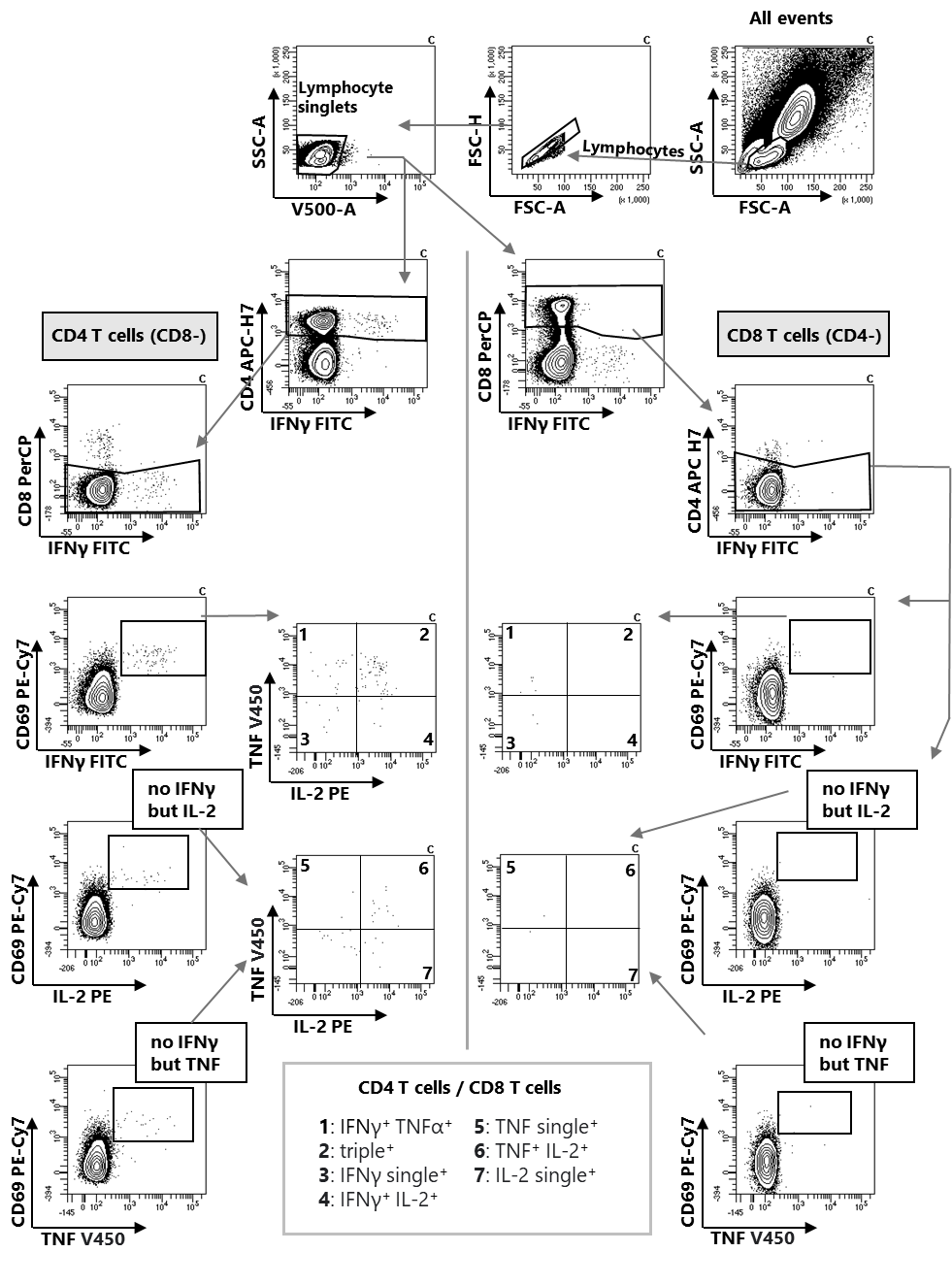

**Supplementary figure S1: Gating strategy for identification of RSV-specific CD4 and CD8 T-cells after stimulation.** Lymphocytes were identified based on cell size (FSC) and granularity (SSC). Doublet elimination was realized by adjusting height and area signals. Blank channel was subtracted. CD4 and CD8 T-cell subpopulations were defined as CD4-positive and CD8-negative or CD4-negative and CD8-positive cells, respectively. CD69-positive cells were selected and further separated according to their cytokine expression using IFNγ, IL-2 or TNF. Boxes were used as gates to quantify the percentage of CD69^+^ CD4 or CD8 T-cells co-expressing a cytokines among all CD4 T-cells.

#### Supplementary figure S2

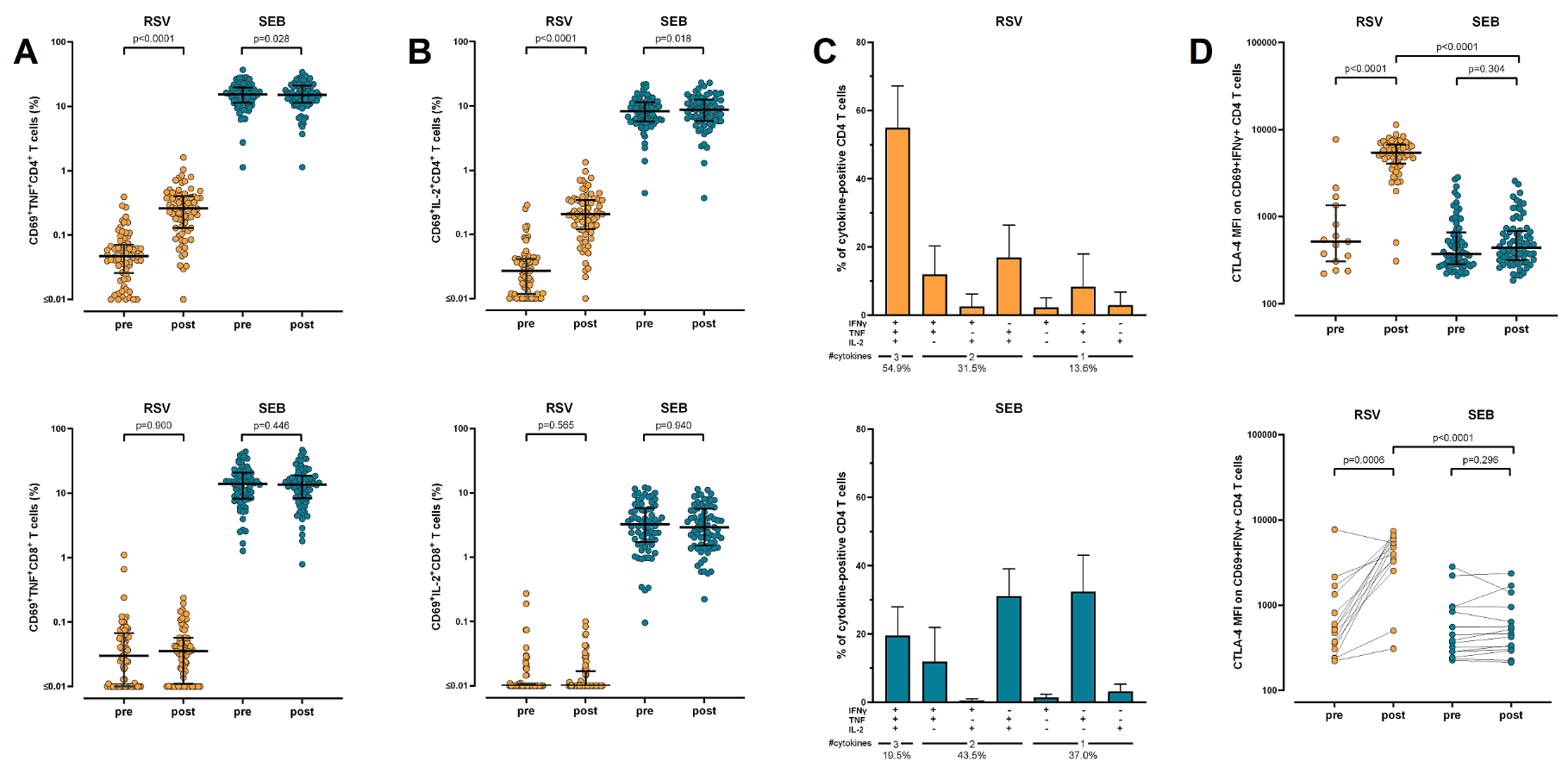

**Supplementary figure S2: Functional and phenotypical characterization of RSV-specific cellular vaccine response in patients with CKD. (A)** Percentages of RSV-specific CD69^+^TNF^+^, and of RSV-specific CD69^+^IL2^+^ **(B)** CD4 and CD8 T-cells before (pre) and 14 days after (post) vaccination, including polyclonal stimulation with SEB. Dots represent individual patients; lines represent medians and interquartile ranges; **(C)** Cytokine profiles of RSV-specific and SEB-reactive CD4 T-cells were calculated defined as percentage of expressing three cytokines (IFNγ, TNF and IL2), two cytokines or a single cytokine only; all samples (from all individuals) were analyzed, but only samples with at least 30 cytokine-expressing CD4 T-cells after normalization to the negative control stimulation were considered to ensure robust statistics (n=61 patients for RSV-specific T-cells, n=72 patients for SEB-reactive T-cells). **(D)** RSV-specific and SEB-reactive CD4 T-cells were analyzed for expression of CTLA-4, which is expressed as median fluorescence intensity (MFI). All samples (from all individuals) were analyzed, but this analysis was restricted to samples with at least 20 CD69^+^IFNγ^+^ CD4 T-cells to ensure robust statistics (15 patients pre, 60 patients post for RSV-specific T-cells, 70 patients pre, 72 patients post for SEB-reactive T-cells). Lines represent medians with interquartile ranges. Differences were calculated using the Mann Whitney test. Moreover, induction of CTLA-4 expression on RSV- and SEB-reactive CD4 T-cells from 15 individuals with detectable RSV-specific CD4 T-cells at baseline are shown before and after vaccination. Differences were calculated using Wilcoxon matched pairs test. CD, cluster of differentiation; CTLA-4, cytotoxic T-lymphocyte antigen 4; IFN, interferon; IL, interleukin, RSV, respiratory syncytial virus; SEB, *Staphylococcus aureus* Enterotoxin B; TNF, tumor necrosis factor.

#### Supplementary Figure S3

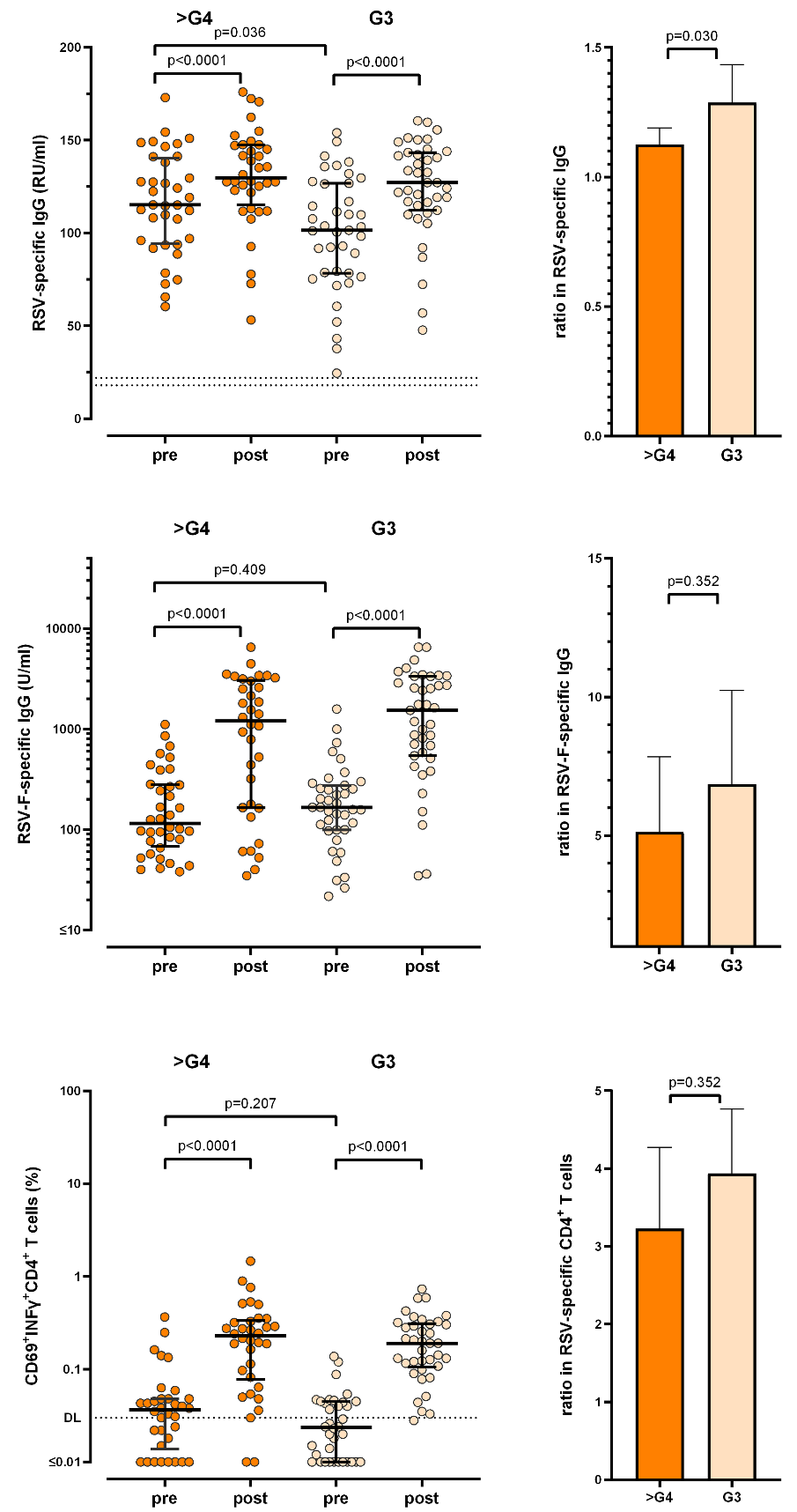

**Supplementary figure S3: Vaccine-induced humoral and cellular immune response in advanced (>G4) vs non-advanced (G3) CKD.** Levels of pan-RSV-specific IgG and RSV-F-IgG, as well as percentages of RSV-specific CD4 T-cells before (pre) and 14 days after (post) vaccination in patients with KDIGO CKD stages G3 (G3a/G3b, n=39) versus CKD stages >4 (n=36), including comparison of the fold increase upon vaccination. Dots represent individual patients; lines represent medians and interquartile ranges; columns represent geometric means with 95% confidence intervals; p-values were calculated from paired values using the Wilcoxon matched pairs test, p-values for fold-increases were calculated using the Mann-Whitney test.

#### Supplementary Figure S4

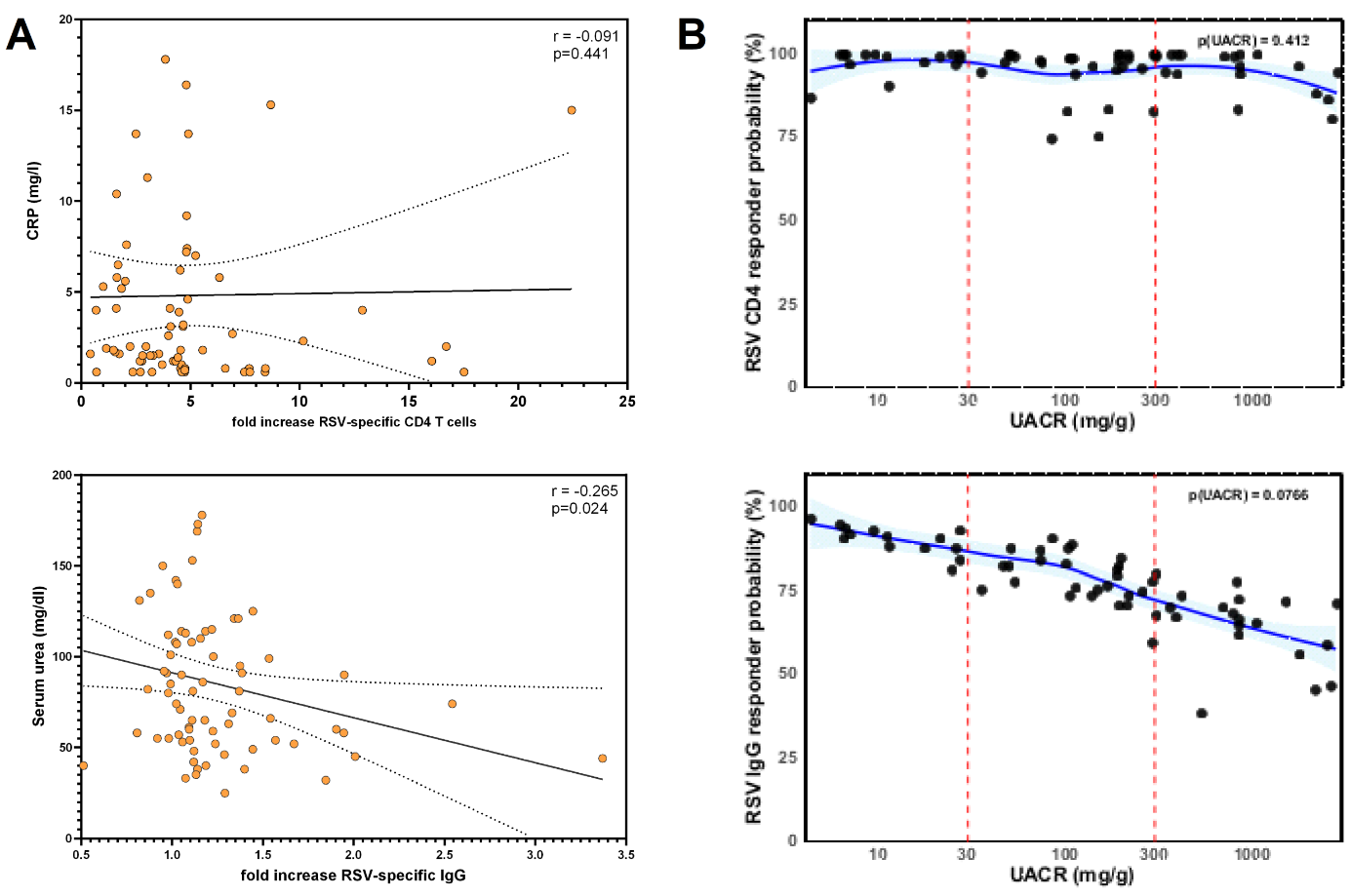

**Supplementary figure S4: Correlations of humoral and cellular vaccine response with clinical CKD parameters pre vaccination and logistic regression modelling of UACR as predictor for vaccine response. (A)** Linear inverse correlations between the fold increase in RSV-specific CD4 T-cells and CRP or RSV-specific IgG and the serum urea. Dots represent individual patients. Lines correspond to the linear regression line with overlaid confidence intervals. **(B)** Predicted probability of being a vaccine responder defined as having an increase in either RSV-specific CD4 T-cells or IgG, based on a multivariate logistic regression model including log-transformed UACR as predictor and eGFR, serum urea and age as covariates. The blue line represents a locally weighted scatterplot smoothing (LOESS) curve with other covariates set at the median, the light blue area displays the 95% confidence interval. Dots represent individual patients. CD, cluster of differentiation; CRP, C-reactive protein; eGFR, estimated glomerular filtration rate; Ig, immunoglobulin; RSV, respiratory syncytial virus; UACR, urine albumin-to-creatinine ratio.
